# First Real-World Evidence Utilizing the Multidimensional Tinnitus Functional Index to Assess Treatment Impact with Bimodal Neuromodulation

**DOI:** 10.64898/2026.01.20.26344445

**Authors:** Miles Sabine, Brian Fligor

## Abstract

**Purpose:** Real-world evidence (RWE) is of practical significance as it enables the evaluation of whether findings observed in rigorously controlled clinical trial settings are generalizable to routine clinical practice. While Lenire, a bimodal neuromodulation tinnitus treatment device, has demonstrated efficacy and safety within controlled trials, further RWE from clinics is needed to reinforce these results. This is the first real-world study to assess the therapeutic effects of Lenire on tinnitus using the Tinnitus Functional Index (TFI), a multidimensional instrument designed to capture tinnitus severity and treatment responsiveness. The study correlates findings with the Tinnitus Handicap Inventory (THI), a well-established tool that assesses the perceived functional, emotional, and catastrophic impact of tinnitus that was used in previous clinical trials and real-world studies. The use of an alternative validated outcome measure in a real-world study may add more feasible, relevant and patient-centered research findings to the body of evidence for Lenire, while maintaining scientific credibility.

**Methods:** A single-site, single-arm retrospective study examining patients fitted with the Lenire device was conducted. Ninety-seven patients with moderate or greater tinnitus severity used the Lenire device for 12 weeks, for up to 60 minutes a day. The primary outcome was change in tinnitus severity, assessed using the TFI at 6-week (FU1) and 12-week (FU2) follow-ups. The THI was included as a secondary outcome measure. Responder rates and mean score changes between initial assessment and FU1 and FU2 were compared using Z-tests for proportions and t-tests, respectively. Pearson’s correlations were used to examine the relationship between the TFI and THI change scores.

**Results:** After just 12 weeks of treatment, 73.4% [95% CI: 62.6%, 84.3%] of patients achieved a clinically significant improvement, defined as a reduction of at least 13 points on the TFI. This improvement was strongly supported by results from the THI, where 84.1% [95% CI: 75.1%, 93.2%] of patients met the minimum clinically important difference of 7 points. Mean score reductions were-25.9 (2.4, SEM) for the TFI and - 28.0 (2.4, SEM) for the THI. Change scores from initial assessment to FU2 on the TFI and THI were highly correlated (r = 0.74, p < 0.001), indicating strong agreement between the two measures in capturing treatment related improvements. All eight TFI subdomains showed reductions ranging from 18.5 to 31.4 points at FU2.

**Conclusions:** This retrospective study demonstrates that 12 weeks of treatment with the Lenire device resulted in clinically meaningful improvements in tinnitus severity on the TFI which was strongly supported by the THI. The high correlation between TFI and THI change scores indicates strong correlation between the two questionnaires in capturing treatment effects. Furthermore, all eight TFI subdomains showed notable reductions, underscoring the multidimensional impact of the treatment. These findings support the clinical utility of both the TFI and THI as complementary tools for evaluating treatment outcomes and guiding tinnitus management in routine practice.

## Introduction

Tinnitus, the perception of sound without an external source, is a prevalent condition affecting an estimated 10–15% of the global population (Heller 2003; Møller et al. 2010; Baguley et al. 2013; McCormack et al. 2016; Biswas and Hall 2020; Biswas et al. 2022). It is often experienced as ringing, buzzing, hissing, or other phantom sounds. While tinnitus may be a minor disturbance for some individuals, it becomes bothersome in an estimated 6–11% of cases (Biswas et al. 2022), with the potential to significantly impair quality of life if left unmanaged. Several therapeutic approaches aim to reduce tinnitus-related distress, and their effectiveness is often evaluated using patient-reported outcome measures. The Tinnitus Functional Index (TFI) provides a multidimensional assessment, encompassing domains such as sleep disturbance, cognitive interference, emotional distress, and auditory difficulties (Meikle et al. 2012), whereas the Tinnitus Handicap Inventory (THI) is widely used to assess the perceived functional, emotional, and catastrophic impact of tinnitus (Newman et al. 1996). Both instruments play a critical role in characterizing tinnitus severity and monitoring treatment outcomes in clinical and research settings.

Lenire® (Neuromod Devices, Dublin, Ireland), a home use bimodal neuromodulation treatment device that combines sound therapy with electrical stimulation of the tongue, has recently gained recognition as an effective tinnitus treatment. Evidence of its effectiveness through preclinical animal research (Markovitz et al. 2015) as well as multiple large-scale clinical trials (Conlon et al. 2020, 2022; Boedts et al. 2024), led to its approval by the Food and Drug Administration (FDA) for the treatment of tinnitus in March 2023 (FDA De Novo request classification number: DEN210033).This approval was largely based on findings from the pivotal, FDA-guided controlled TENT-A3 trial (Boedts et al. 2024). Results demonstrated that participants presenting with bothersome tinnitus symptoms (THI score ≥38) (Newman et al. 1996), experienced clinically meaningful improvements (that is, met or exceeded the Minimal Clinically Important Difference; MCID), marked by a reduction of more than seven points on the THI scale (Zeman et al. 2011). In TENT-A3, outcomes with bimodal neuromodulation significantly outperformed those obtained from sound-only stimulation (Boedts et al. 2024).

Recent real-world evidence (RWE) from a United States clinic further supports the effectiveness of Lenire in everyday clinical practice (McMahan and Lim 2025). A retrospective study conducted at the Alaska Hearing & Tinnitus Center (AHTC) (McMahan and Lim 2025) reported results consistent with those of earlier large-scale clinical trials (Conlon et al. 2020, 2022; Boedts et al. 2024) and prior RWE from European clinics (Buechner et al. 2022; Boedts et al. 2024). At AHTC, after approximately 12 weeks of treatment with Lenire, 91.5% of patients who initially presented with bothersome tinnitus (THI score ≥38) achieved clinically meaningful improvement on the THI scale (McMahan and Lim 2025). As Lenire becomes more widely adopted in clinical practice, the continued collection and analysis of real-world data is essential to further validate its effectiveness, monitor treatment outcomes across diverse populations, and inform best practices for clinical implementation.

To date, clinical trials and RWE studies evaluating the efficacy of the Lenire device have primarily used the THI as the outcome measure. Its consistent use across studies has enabled reliable comparisons across both controlled and real-world settings. The TFI (Meikle et al. 2012) has also been incorporated in key Lenire clinical trials, including TENT-A1 (Conlon et al. 2020) and TENT-A2 (Conlon et al. 2022), where it complemented the THI. However, the TFI has not yet been used in real-world clinical evaluations of Lenire.

Tinnitus treatment outcomes are difficult to quantify using psychophysical measures (e.g., pitch-matching and loudness-matching), as these do not adequately reflect the subjective and often life disrupting impact of the condition. While consistency in outcome measurement using the THI is important for cross-study comparisons, incorporating additional validated instruments, such as the TFI, may provide complementary insights into how patients experience and respond to treatment. This is particularly relevant in clinical practice, where understanding the full scope of tinnitus burden is essential for guiding individualized care. As noted by (Langguth and De Ridder 2023), the TFI offers a broader and more balanced account of tinnitus-related distress, albeit with a higher minimum clinically important difference (MCID) of 13 points compared to the 7-point MCID of the THI.

Including both the TFI and THI in treatment evaluation can strengthen the robustness of clinical findings. The use of multiple validated tools strengthens the reliability of outcome assessments and improves the ability to verify that treatment related changes are accurately captured. To our knowledge, this is the first real-world, single-site, single-arm retrospective chart review in the United States to assess Lenire’s clinical effectiveness using both the TFI and THI. The objectives of this retrospective chart review conducted at Tobias & Battite Hearing Wellness (T&B) were to evaluate the use of the TFI as an outcome measure for Lenire treatment in clinical practice, examine the correlation between TFI and THI outcomes, and determine whether treatment outcomes observed on the THI scale in previous Lenire trials (Conlon et al. 2020, 2022; Boedts et al. 2024) and RWE studies (Buechner et al. 2022; Boedts et al. 2024; McMahan and Lim 2025) can be replicated in a different clinical setting.

## Methods

### Study design and standard of care

This retrospective, single-site, single-arm study examined data from 97 patients with bothersome tinnitus who were assessed and fitted with the Lenire device (**Figure 1a**) between May 19, 2023, and February 29, 2024, at Tobias & Battite Hearing Wellness (T&B). T&B is a privately-owned and operated audiology practice at two locations (Boston and Beverly, Massachusetts) that has been in continuous operation for over 50 years. The authors are the two licensed audiologists who conducted audiological evaluation, tinnitus treatment evaluation, and management recommendations for all patients included in this study. All patients attended an initial in-person assessment with one of the two authors. These initial appointments included the collection of a comprehensive history, including the TFI and THI, audiometry, acoustic immittance, and, where clinically relevant, further specialized audiological testing was conducted. Patients were referred for further evaluation and management with otolaryngology, psychology, psychiatry, and/or other healthcare providers as appropriate. As part of standard clinical care, patients also received tinnitus education and structured counselling at the initial assessment and as appropriate during follow-up appointments. Tinnitus education and counselling were focused on helping the patients better understand the nature of their tinnitus, setting realistic expectations for treatment, and reducing the impact of tinnitus on daily life.

**Figure 1:**
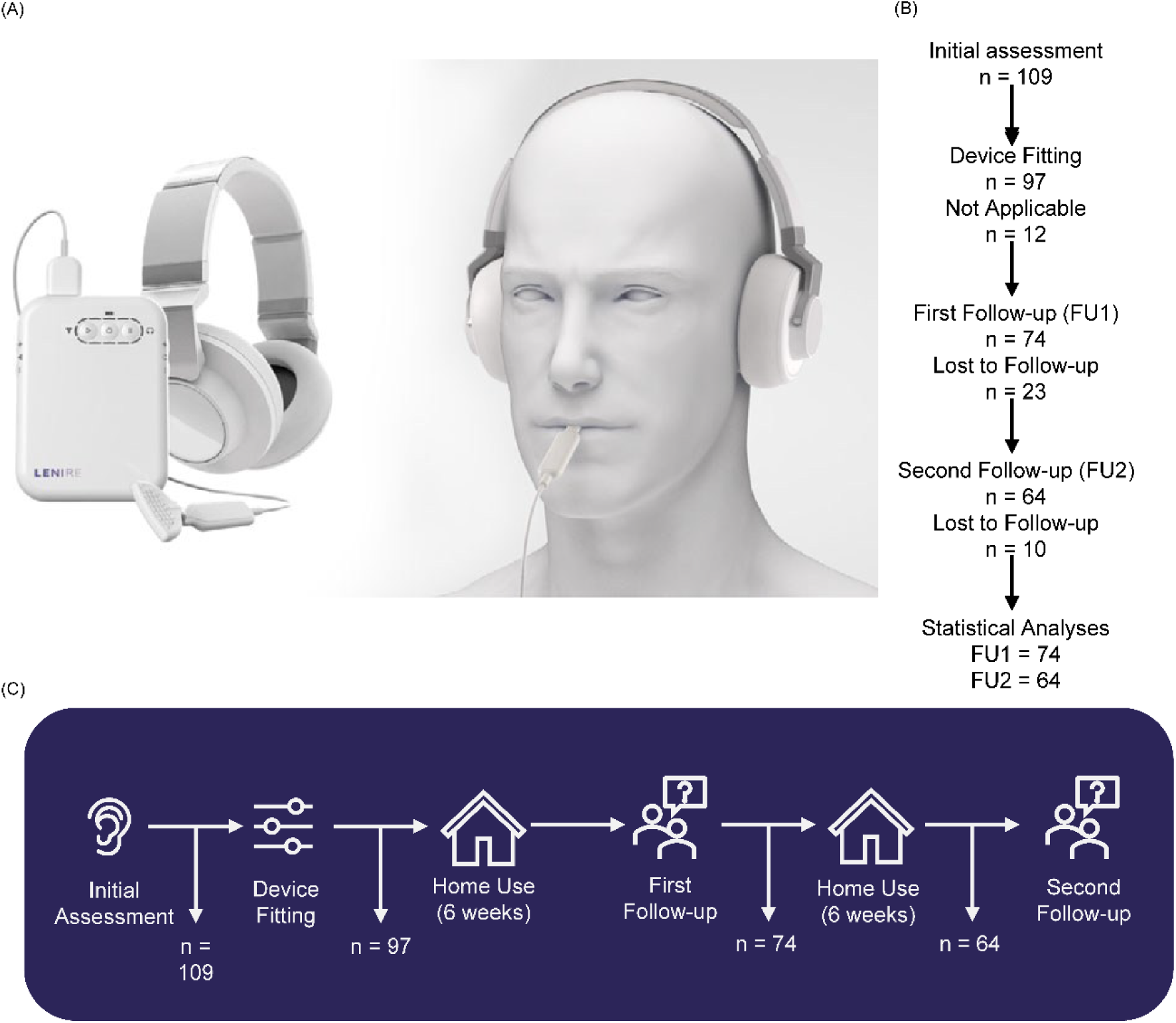
a). The Lenire bimodal neuromodulation device developed by Neuromod Devices (Boedts et al. 2024) is intended to reduce tinnitus symptoms in patients with moderate or greater severity (defined as a Tinnitus Handicap Inventory (THI) score ≥38). The system is comprised of three components: the Tonguetip®, an intraoral device designed to rest comfortably in the mouth and deliver mild electrical stimulation to the surface of the tongue, the Bluetooth headphones, which deliver personalized auditory stimulation, and the handheld controller, which allows patients to adjust the intensity and duration of their treatment sessions. b). Patient disposition per visit for the study’s primary outcome (TFI) among patients who had at least moderate tinnitus severity (THI ≥38). c). Lenire standard of care procedure at Tobias and Battite Hearing Wellness (T&B).

Towards the end of the initial assessment patients were informed of the treatment options they were eligible for including Lenire, hearing aids, tinnitus masking, tinnitus retraining therapy and referral to licensed mental health counselors. Patients opting to proceed with Lenire treatment were fitted with the device either during the initial visit or at a separate in-person appointment. The audiologist provided hands-on training, reviewed the Lenire User Manual with the patient, and calibrated the device to each patient’s needs by setting the tongue stimulus to a comfortable level and adjusting the auditory stimulus based on the patient’s audiogram. The audiologist also discussed expected outcomes and emphasized the importance of treatment adherence.

Lenire is an FDA-approved, at-home neuromodulation device for the treatment of tinnitus. It delivers pure tones through headphones, synchronized with mild electrical pulses to the tongue via the Tonguetip®. Treatment is self-administered by patients at home, with instructions to use the device for 60 minutes daily over a 12-week period. Two stimulation settings, PS1 and PS6, are FDA-approved based on findings from the TENT-A3 pivotal trial and additional real-world data included in the FDA’s De Novo submission. In this study, all patients used the PS1 setting. Detailed descriptions of PS1 and PS6 are available in prior publications (Conlon et al. 2020, 2022; Boedts et al. 2024).

Follow-up assessments were conducted approximately six weeks (Follow-up 1; FU1) and 12 weeks (Follow-up 2; FU2) after device fitting (**Figure 1b**, **Figure 1c**). Tinnitus severity was reassessed at each visit using the TFI and THI. In-person visits were the preferred format for follow-up assessments and consultations. However, virtual video calls were offered as an alternative when needed. Patients were also encouraged to contact the clinic between appointments if any issues arose.

Data from these clinic visits, including audiological assessments and patient-reported outcomes, were extracted from the patients’ electronic medical records, which were maintained in the CounselEar Office Management System (OMS). All data were de-identified before being included in the study database, with no re-identification required.

All patients had previously provided written consent to a HIPAA Waiver Notice (“Notice of Privacy Practice”) during their initial clinic visits, allowing their clinical data to be used for research purposes. One patient requested their data to be excluded from analysis. The specific intention to use data for Lenire-related research was explained verbally during initial assessment, and patients provided informed consent accordingly.

The study protocol was reviewed by Advarra Institutional Review Board (IRB number: 00000971; Protocol number: Pro00079438) and was deemed exempt from full IRB review in accordance with the United States Department of Health and Human Services regulations (45 CFR 46.104(d)(4)).

### Indications and contraindications for Lenire use

Lenire is intended for adults aged 18 and older who experience subjective tinnitus with moderate or worse severity (THI ≥38). It should not be used unless specifically advised by a physician or dentist in individuals who are pregnant, have an active implantable device, or have reduced tongue sensitivity. It is also contraindicated for individuals with epilepsy or other conditions that may lead to loss of consciousness, as well as those with sores, lesions, or inflammation in the oral cavity, chronic or intermittent neuralgia in the head and neck, or Meniere’s disease, unless approved by a physician. Additionally, Lenire should not be used by individuals with objective tinnitus only or those unable to remove oral piercings during device use.

### Assessments

This study aimed to explore the treatment response to Lenire using the TFI alongside the more widely reported THI in order to compare treatment outcomes between the two questionnaires. The TFI (Meikle et al. 2012) is a validated questionnaire consisting of 25 questions that assesses the patient’s tinnitus experience over the previous week. Each item is scored on a scale from 0 to 10, or 0%–100%, with higher scores indicating greater tinnitus severity. The questionnaire is divided into eight subscales of tinnitus impact: Intrusiveness, Sense of Control, Cognitive, Sleep, Auditory, Relaxation, Quality of Life, and Emotional. To be considered valid, a completed TFI questionnaire must have responses to at least 19 out of the 25 items, with no more than one question omitted per subscale. The subscale score per patient was calculated as the summation of their possible subscale score, divided by the number of possible answers, and multiplied by 10 to obtain a value from 0 to 100. A total higher score reflects a greater negative impact of tinnitus on the patient’s life. While the TFI was originally developed by (Meikle et al. 2012), (Henry et al. 2016) later proposed a five-level problem classification system to aid in interpreting the global score of the TFI: 0-17 (not a problem), 18-31 (small problem), 32-53 (moderate problem), 57-72 (big problem), 73-100 (very big problem).

Similar to the TFI, the THI (Newman et al. 1996) is a validated 25-item questionnaire designed to assess the severity of tinnitus and its impact on a patient’s daily life. Each question is answered with one of three options: “Yes” (scored as 4), “Sometimes” (scored as 2), or “No” (scored as 0). The total score is calculated by summing the individual responses, resulting in a score range from 0 to 100, with higher scores indicating a greater level of tinnitus-related handicap. The THI categorizes tinnitus severity into five levels based on the total score: 0–16 (slight), 18–36 (mild), 38–56 (moderate), 58–76 (severe), and 78–100 (catastrophic). The questionnaire is designed to assess the impact of tinnitus across three main domains: functional, emotional, and catastrophic.

According to (Langguth and De Ridder 2023), the THI and TFI assess overlapping but distinct dimensions of tinnitus-related distress. The THI places greater emphasis on the emotional and psychological impact of tinnitus, while the TFI encompasses a broader range of functional domains, including sleep, auditory perception, and specific tinnitus characteristics such as perceived intrusiveness and annoyance (Langguth and De Ridder 2023). As such, the THI and TFI offer complementary perspectives on the multidimensional experience of tinnitus and can be used together to achieve a more comprehensive assessment.

### Field safety reporting

Healthcare professionals can report adverse events (AEs), technical issues, or general feedback directly to the manufacturer, Neuromod Devices, via the Zoho ticketing system. Each submission is assessed for potential device malfunctions, fitting issues, or undesirable medical symptoms. If warranted, a Field Product Experience Report (FPER) is initiated and reviewed by the technical team to determine if regulatory reporting to the FDA (in the United States) or Competent Authorities (outside the United States) is required. In this study, one medical FPER was submitted due to patient discomfort during tongue stimulation. All other technical queries were resolved without the need for regulatory reporting.

### Data and Statistical Analysis

“Responder rates” were calculated for both the TFI and THI as the percentage of patients whose scores improved by at least the established MCID: 13 points for the TFI (Meikle et al. 2012) and 7 points for the THI (Zeman et al. 2011). These responder rates were assessed from initial assessment to follow-up one (FU1, at 6 weeks post-treatment initiation) and follow-up two (FU2, at 12 weeks post-treatment initiation). Corresponding 95% Confidence Intervals were reported for each time point. To explore the robustness of these outcomes, additional responder analyses using alternative thresholds, 15% improvement (Langguth and De Ridder 2023) and 9-point reduction (Engelke et al. 2025) for TFI, and 15% improvement (Langguth and De Ridder 2023) and 11-point reduction (Engelke et al. 2025) for THI are reported in **Supplementary Figure 1**. Mean score changes from initial assessment to each follow-up were also calculated for both TFI and THI measures and presented with standard error of the mean (SEM).

To compare the responder rates between the initial assessment and each follow-up (FU1 and FU2), two-sided z-tests of proportions were conducted. Differences in mean score changes between the initial assessment and each follow-up were evaluated using independent t-tests. Pearson’s correlation coefficients were calculated to examine the relationship between the TFI and THI at initial assessment, as well as between their respective change scores.

To account for missing data for the primary (TFI) and secondary (THI) endpoints resulting from drop-out rates of 23.7% at FU1 and 34% at FU2, multiple imputation was conducted as a sensitivity analysis using the mice package (version 3.17) in R (version 4.4.3). The imputation employed Markov Chain Monte Carlo (MCMC) predictive mean matching (PMM) to generate plausible values for missing follow-up TFI and THI scores. Predictor variables included TFI and THI scores, age, gender, and tinnitus duration at initial assessment to improve the accuracy of imputed values. The imputation procedure consisted of 50 iterations to ensure convergence and robustness of the imputations. Post-imputations, analyses were repeated and showed minimal differences compared to complete-case results as detailed in **Supplementary Table 1.** All analyses were performed using R (version 4.4.3) with p-values less than 0.05 considered statistically significant.Error! Reference source not found.

### Data quality assurance

The primary analysis database, which included TFI and THI scores, was verified against the source data (CounselEar) during an on-site monitoring visit. Source Data Verification (SDV) was performed for 100% of the data for all patients in this study. This comprehensive approach was adopted due to the manual transcription of source data by clinicians, which carries a risk of entry errors. Any discrepancies found were corrected during the visit for THI and TFI. The analysis was conducted on the final validated dataset. Monitoring was carried out by qualified, trained personnel who were briefed beforehand and were independent of the investigation site. The monitoring took place on May 20, 2024.

## Results

### Patient disposition and characteristics

A total of 97 patients with moderate and above tinnitus severity (THI ≥38) with available TFI scores at initial assessment were fitted with the Lenire device between May 19, 2023, and February 29, 2024 (**Figure 1b**). This threshold reflects the clinical indication for Lenire, which is intended for patients with a THI score of 38 or higher. Of the 97 patients, 76.2% (n=74) returned for FU1 and 65.9% (n=64) returned for FU2. Regarding the secondary outcome measure, all patients completed the THI questionnaire at initial assessment. However, eight patients at FU1 and one patient at FU2 did not complete the THI, resulting in 66 and 63 patients completing THI data at these visits, respectively. The age of the patients at the time of the initial assessment ranged from 21 to 81, with a mean of 58.5 years (12.4, SD; **Table 1**). The cohort consisted of 68.0% males (n=66) and 32.0% females (n=31; **Table 1**). The mean initial TFI score was 59.3 points (15.2, SD), while the mean THI score was 60.4 points (16.5, SD; **Table 1**). The patient’s tinnitus duration at the time of device fitting was categorized as follows: less than one year (15.5%, n=15), 1 to under 5 years (44.3%, n=43), 5 to under 10 years (17.5%, n=17), 10 to under 20 years (13.4%, n=13) and greater than or equal to 20 years (9.3%, n=9; **Table 1**).

**Table 1:** Demographic characteristics of 97 patients who were fitted with the Lenire device with TFI scores.

| Characteristic | Value |
| --- | --- |
| Age (years) Mean ± SD (n) | 58.5 ± 12.4 (97) |
| Gender [% (n/N)] |  |
| Male | 68% (66/97) |
| Female | 32% (31/97) |
| TFI at initial assessment (points) |  |
| Mean ± SD (n) | 59.3 ± 15.2 (97) |
| THI at initial assessment (points) |  |
| Mean ± SD (n) | 60.4 ± 16.5 (97) |
| Tinnitus duration (years) [% (n/N)] |  |
| <1 | 15.5% (15/97) |
| 1 to <5 | 44.3% (43/97) |
| 5 to <10 | 17.5% (17/97) |
| 10 to <20 | 13.4% (13/97) |
| ≥20 | 9.3% (9/97) |

### High clinically meaningful response on the TFI scale

In this study, 73.4% [95% CI: 62.6%, 84.3%] of patients with moderate and above tinnitus achieved the MCID of at least a 13-point reduction on the TFI after 12 weeks of treatment (FU2; **Figure 2a**). This corresponded to a mean reduction of 25.9 (2.4, SEM) points in tinnitus severity (**Figure 2b**). Encouragingly, a substantial proportion of patients, 70.3% [95% CI: 59.9%, 80.7%] (**Figure 2a)**, already demonstrated clinically meaningful improvement after approximately six weeks of treatment (FU1), with a mean TFI reduction of 22.1 (2.0, SEM) points (**Figure 2b**). There were no statistically significant differences in proportion of responders (p = 0.68) (**Figure 2a)** or in the mean TFI change (p = 0.225) (**Figure 2b)** between initial assessment and FU1 and initial assessment and FU2, suggesting that therapeutic effects were achieved early and then maintained.

**Figure 2:**
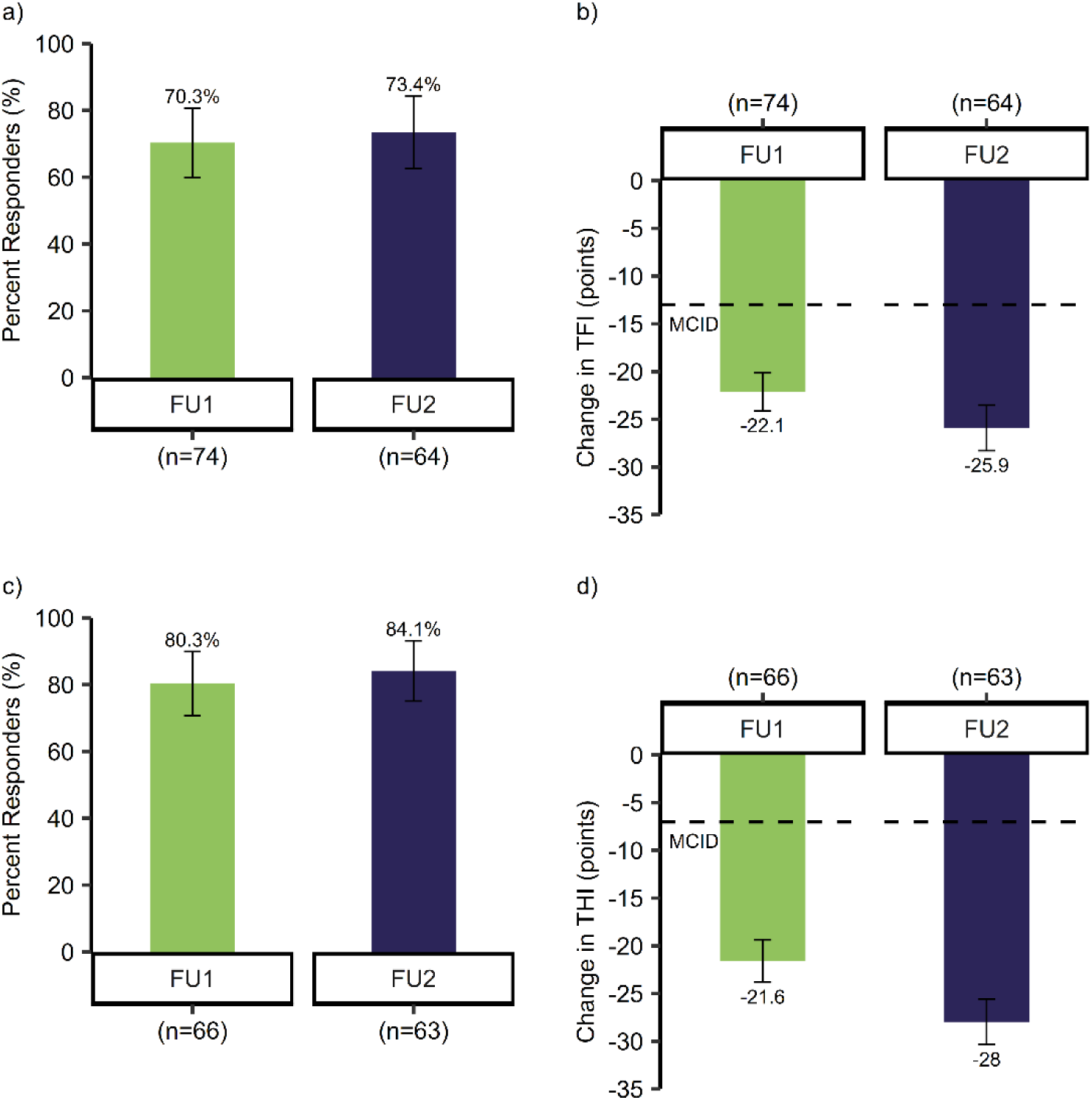
a) Percent responders using change in TFI greater than the MCID of-13 points. 70.3% [95% CI: 59.9%, 80.7%] at FU1 (n=74) and 73.4% [95% CI: 62.6%, 84.3%] at FU2 (n=64). Two-sided z-test for proportions, p-value of 0.68. b) Mean change in TFI from initial assessment with a decrease of-22.1 (2.0, SEM) TFI points at FU1 (n=74) and-25.9 (2.4, SEM) TFI points at FU2 (n=64) with a p-value between timepoints of 0.225. c) Percent responders using change in THI greater than the MCID of-7 points. 80.3% [95% CI: 70.7%, 89.9%] at FU1 (n=66) and 84.1% [95% CI: 75.1%, 93.2%] at FU2 (n=63). Two-sided z-test for proportions, p-value of 0.571. d) Mean change in THI from initial assessment with a decrease of-21.6 (2.2, SEM) points at FU1 (n=66) and-28.0 (2.4, SEM) points at FU2 (n=63) with a p-value between timepoints of 0.053.

Using the alternative MCID threshold of a 9-point reduction, 77.0% [95% CI: 67.4%, 86.6%] and 78.1% [95% CI: 68.0%, 88.3%] of patients were classified as responders at FU1 and at FU2, respectively (**Supplementary Figure 1a**). Similarly, when defining response as a greater than 15% reduction in TFI score, 78.4% [95% CI: 69.0%, 87.8%] of patients showed improvement at FU1, and 76.6% [95% CI: 66.2%, 86.9%] showed improvement at FU2 (**Supplementary Figure 1b**). *Figure 2Figure 2*

### THI results reinforce significant tinnitus improvement

Furthermore, our THI results support the TFI findings, with 84.1% [95% CI: 75.1%, 93.2%] of patients exceeding the MCID of a 7-point reduction at FU2 (**Figure 2c**). This corresponded to a mean 28.0 (2.4, SEM) point reduction in tinnitus severity (**Figure 2d**). Clinically significant improvement was also achieved by 80.3% [95% CI: 70.7%, 89.9%] of patients at FU1 (**Figure 2c**) with a mean reduction of 21.6 (2.2, SEM) points in THI scores (**Figure 2d**). Statistical comparisons showed no significant differences in either the proportion of responder (p = 0.571) (**Figure 2c**) or the mean score reduction (p = 0.053) (**Figure 2d**) between initial assessment and FU1 and initial assessment and FU2. Similar to TFI results, THI findings suggest that the majority of treatment benefits were achieved in the first 6 weeks and sustained through 12 weeks.

When applying a stricter MCID threshold of 11 points, responder rates were 71.2% [95% CI: 60.3%, 82.1%] at FU1 and 79.4% [95% CI: 69.4%, 89.4%] at FU2 (**Supplementary Figure 1c**). Similarly, when considering a response as > 15% reduction in THI scores, 77.3% [95% CI: 67.2%, 87.4%] of patients showed improvement at FU1 and 84.1% [95% CI: 75.1%, 93.2%] at FU2 (**Supplementary Figure 1d**). These results highlight the extent and robustness of treatment benefit.

### Movement toward lower tinnitus severity across THI and TFI categories

Because THI scores correspond to defined severity categories, analyzing shifts between these categories over time provides additional insight into treatment outcomes. At FU2, there was a marked reduction in patients classified within the higher severity categories (**Supplementary Figure 2a**). Specifically, the proportion of patients in the ‘catastrophic’ category dropped from 20.4% at initial assessment to 0% at FU2. The ‘severe’ group similarly declined from 26.5% to 2.0%, while those categorized as ‘moderate’ decreased from 53.1% to 22.5%. These results illustrate a clear overall movement toward milder tinnitus severity. While THI categories are widely adopted, TFI severity categories, such as those proposed by (Henry et al. 2016) are increasingly used in clinical and research settings. A similar pattern of improvement was observed when applying these TFI severity categories, as shown in **Supplementary Figure 2b**.

### TFI and THI scores align in measuring the benefits of Lenire treatment

At initial assessment, there was a strong positive correlation between THI and TFI scores (r=0.68, p<0.001, n=97; **Figure 3a**). Of the 97 patients with bothersome tinnitus (THI ≥ 38), only 3 patients were classified below the moderate problem cut-off (TFI < 32) on the TFI scale (**Figure 3a**). The correlation between changes in scores from initial assessment to FU1 was similarly strong (r=0.71, p<0.001, n=66; **Figure 3b**). This relationship remained consistent from initial assessment to FU2 (r=0.74, p<0.001, n=63; **Figure 3c**). These results demonstrate a robust and statistically significant association between THI and TFI measurements throughout the course of treatment.

**Figure 3:**
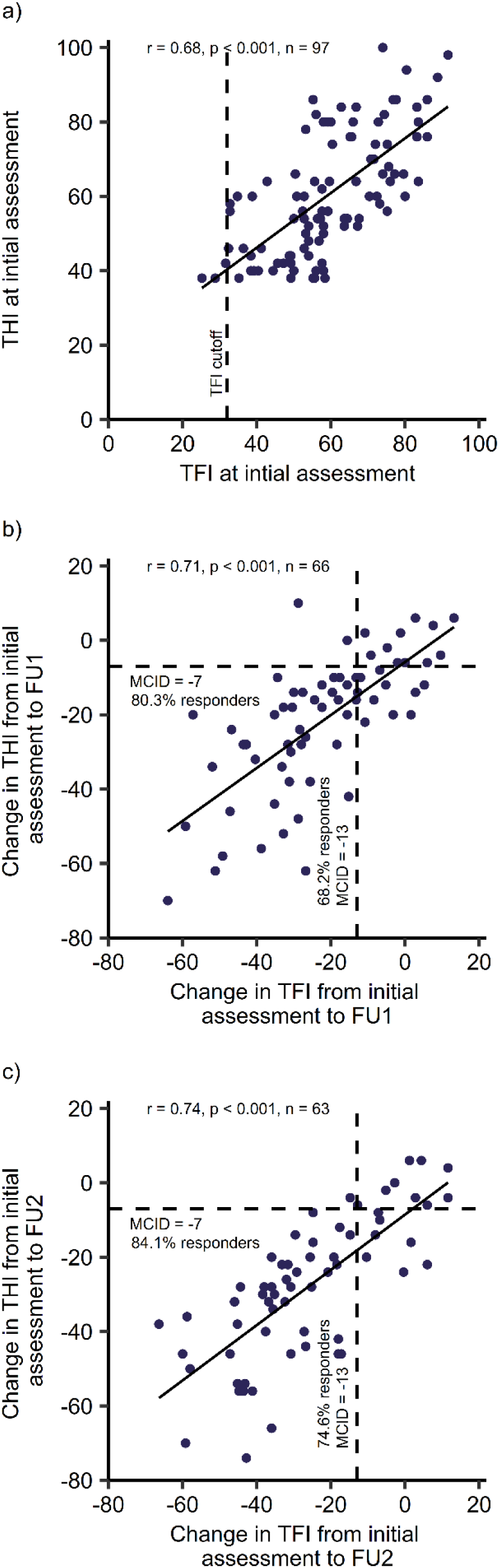
a) Correlation between TFI and THI at initial assessments, r=0.68, p<0.001, n=97. b) Correlation between the changes in TFI and THI from initial assessment to FU1, r=0.71, p<0.001, n=66. 68.2% responder rate for TFI and 80.3% responder rate for THI. c) Correlation between the changes in TFI and THI from initial assessment to FU2, r=0.74, p < 0.001, n=63. 74.6% are responders of TFI, with 84.1% responders of THI.

### Multidimensional symptom relief evident with TFI and THI subscales

The TFI and THI subscales were analyzed to provide a more nuanced understanding of treatment effects at FU2. While the TFI is designed to be multidimensional, capturing various aspects of tinnitus-related distress, the THI is more limited in scope (Langguth and De Ridder 2023). Nonetheless, we included a subscale analysis of the THI to determine whether it could offer complimentary insights. At FU2, substantial mean improvements (**Figure 4a**) and large effect sizes (**Figure 4b**) were observed across all TFI subscales: Intrusive (-26.2 (2.6, SEM), d = 1.28 [95% CI: 0.95, 1.61]), Sense of Control (-21.7 (3.3, SEM), d = 0.81 [95% CI: 0.53, 1.09]), Cognitive (-23.0 (2.6, SEM), d = 1.10 [95% CI: 0.79, 1.41), Sleep (-28.3 (3.2, SEM), d = 1.12 [95% CI: 0.81, 1.43]), Auditory (-18.5 (2.9, SEM), d = 0.79 [95% CI: 0.51, 1.07]), Relaxation (-31.4 (3.5, SEM), d = 1.13 [95% CI: 0.82, 1.44]), Quality of Life (-28.9 (3.0, SEM), d = 1.21 [95% CI: 0.89,1.53), and Emotional (-28.3 (3.9, SEM), d = 0.91 [95% CI: 0.62, 1.20]). Since the TFI subscales are scored differently and do not share a common scale, effect sizes provide a standardized way to compare the magnitude of change across them. While specific cut-off values for each TFI subscale have not been established, these results suggest that Lenire notably reduces the intrusiveness of tinnitus but has substantial impact across all domains highlighting the functional impact provided by Lenire.

**Figure 4:**
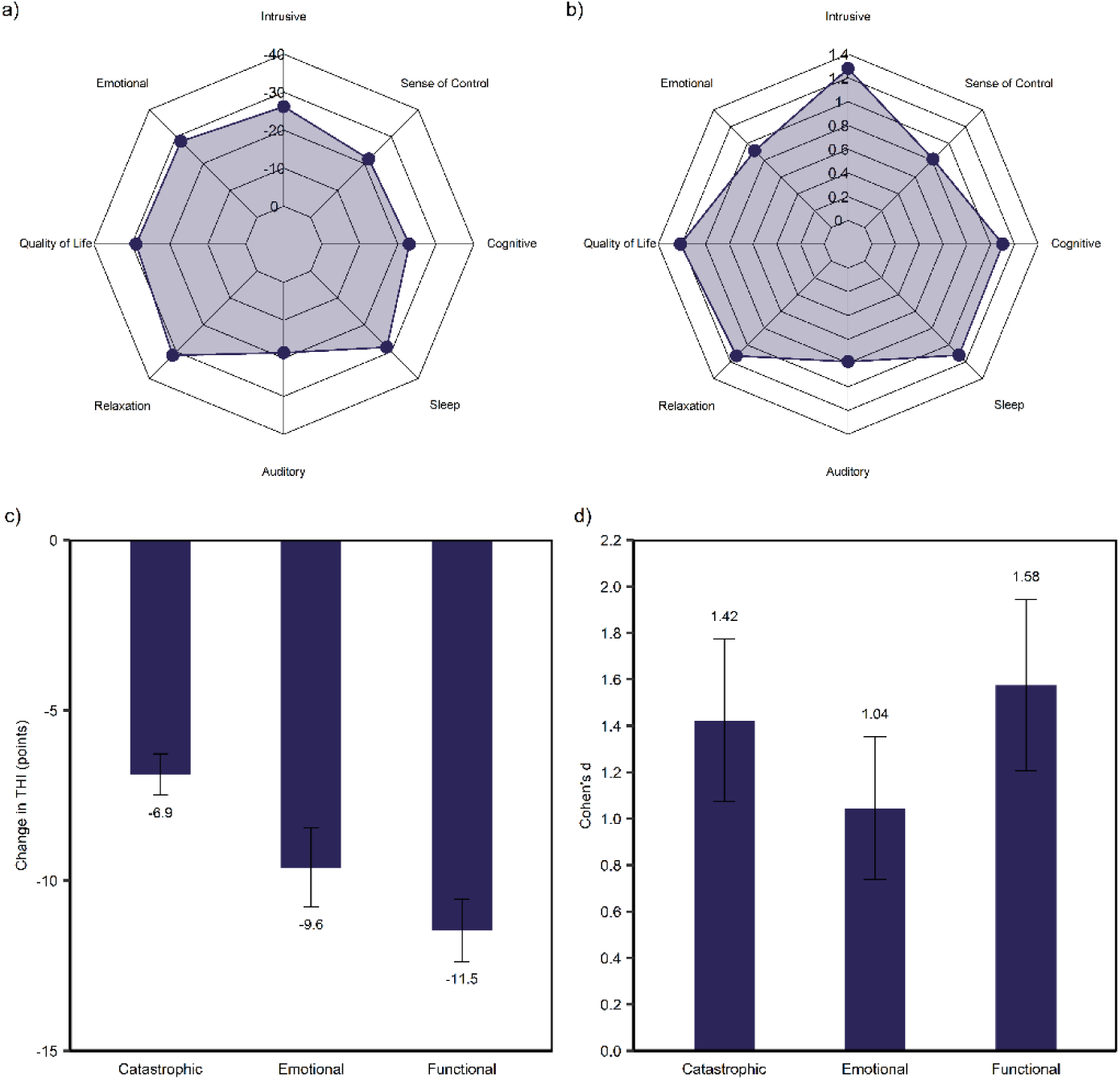
Subscale breakdown from initial assessment to FU2 by tinnitus assessment. a) Mean changes between timepoints across TFI subscales, (n=64). b) Effect sizes between timepoints across TFI subscales, (n=64). c) Mean changes between timepoints across THI subscales, (n=63). d) Effect sizes between timepoints across THI subscales, (n=63).

Similarly, the THI subscales demonstrated notable large mean reductions at FU2 **(Figure 4c)** with large effect sizes (**Figure 4d**): Catastrophic (-6.9 (0.6, SEM), d = 1.42 [95% CI: 1.07, 1.77]), Emotional (-9.6 (1.2, SEM), d = 1.04 [95% CI: 0.74, 1.35]), and Functional (-11.5 (0.9, SEM), d = 1.58 [95% CI: 1.21, 1.94]).

## Discussion

The efficacy of bimodal neuromodulation using combined sound therapy and electrical tongue stimulation for tinnitus treatment has been well established through several large-scale clinical trials (Conlon et al. 2020, 2022; Boedts et al. 2024), including a pivotal controlled study that supported Lenire’s FDA De Novo approval (Boedts et al. 2024). Beyond these controlled clinical trial settings, published RWE from Europe (Buechner et al. 2022; Boedts et al. 2024) and, more recently, from a clinical practice in the United States (McMahan and Lim 2025) further support Lenire’s effectiveness in treating bothersome tinnitus. The current study not only adds to this body of RWE but, for the first time, demonstrates a strong relationship between two widely used tinnitus measurement tools, the THI and TFI, in detecting treatment-related changes after a Lenire intervention in clinic. These findings suggest meaningful convergence between these two commonly used tinnitus measures in real-world practice.

These consistent findings across both clinical trials and real-world settings reinforce the clinical utility of Lenire for patients experiencing moderate to more severe tinnitus. Together, this body of evidence highlights Lenire as a valuable treatment option that delivers significant and meaningful benefits to patients within this indicated population.

As the adoption of Lenire expands, continuous collection and analysis of real-world data is essential to further confirm its effectiveness, assess treatment outcomes across varied patient populations, and inform best practices for clinical application. Previous RWE studies have used THI to measure treatment response for Lenire (Buechner et al. 2022; Boedts et al. 2024), but the equally validated TFI offers complementary insights. Using both tools can strengthen outcome reliability. To our knowledge, this is the first RWE study to assess the effectiveness of Lenire using both the TFI and THI. Results indicate that 73.4% of 64 patients with bothersome tinnitus achieved a clinically meaningful improvement (MCID) on the TFI (Meikle et al. 2012) after approximately 12 weeks of Lenire treatment. The robustness of our TFI findings is reinforced by applying alternative thresholds for responder criteria. When using a 9-point MCID (Engelke et al. 2025), 78.1% of patients were classified as responders at FU2. Similarly, defining response as a greater than 15% reduction (Langguth and De Ridder 2023) in TFI scores yielded comparable improvements, highlighting the consistency of positive outcomes across different criteria.

The TFI changes observed herein are consistent with the THI reductions achieved in this population. Specifically, 84.1% of patients exceeded the MCID threshold on the THI scale (Zeman et al. 2011) at FU2. Responder rates based on alternative MCID thresholds, e.g., greater than a 15% or an 11-point decrease in THI, were similar. These consistent findings highlight the robust therapeutic effects of Lenire across both the TFI and THI. These results showcase that the treatment benefit of Lenire is not restricted to the slightly narrower focus of the THI (i.e. intrusive, emotional and impact on daily life) but comparable across the same and additional dimensions addressed in the TFI across a higher resolution scale. Furthermore, the strong correlations observed between the TFI and THI across timepoints support their consistent use in assessing Lenire treatment outcomes and suggest convergent validity in a real-world clinical context. The categorical shifts in tinnitus severity levels provide further clinical context for the observed improvements. By FU2, there was a marked reduction in proportion of patients classified within the ‘catastrophic’, ‘severe’, and ‘moderate’ THI categories, consistent with a previous RWE study (McMahan and Lim 2025). Importantly, similar pattern of improvement was observed when applying TFI severity categories, as defined by (Henry et al. 2016), reinforcing the robustness of the results across two widely used measurement tools. These observed categorical improvements are clinically meaningful, as lower tinnitus severity is generally associated with minimal daily interference, while higher severity often impacts sleep, quality of life, and mental health (McCombe et al. 2001). Taken together, these results suggest that Lenire may lead to tangible, multidimensional improvements in tinnitus-related distress and overall well-being.

While the THI is traditionally reported using a three-category scoring system that has been validated (Newman et al. 1996), there are ongoing discussions within the scientific community advocating for its analysis as an ordinal response to better capture gradations in severity (Tyler et al. 2006, 2007). In this study, we reported the THI as a continuous and categorical measure. Our results demonstrate that when treated as a continuous variable, the THI correlates well with the TFI, which is measured on a 0–10 scale. By utilizing a scale that has a higher resolution than the THI, it can be reinforced that positive improvements reported via the THI are not inflated due to its lower response resolution. This alignment further supports the validity of the THI in capturing changes in tinnitus severity.

Further examination of the TFI subscales provides a more nuanced understanding of the specific ways in which tinnitus affects patients at T&B, beyond the overall severity score. Analysis of these subscales’ outcomes enables clinicians to identify specific domains of tinnitus-related distress, thereby facilitating more tailored and effective patient care. In this study, all TFI subscales showed substantial improvement at FU2, with particularly large mean reductions observed in the Relaxation, Quality of Life, Emotional, and Sleep domains (each exceeding at least a 28-point reduction). Effect sizes across these subscales were consistently large (all > 0.79), with the Intrusiveness subscale showing the most pronounced effect (d = 1.28). Given that Intrusiveness reflects the persistence, unpleasantness, and disruptive nature of tinnitus, this finding suggests that Lenire may be especially effective in reducing the most distressing aspects of the condition. Since the TFI subscales are scored differently and do not share a common scale, effect sizes provide a standardized way to compare the magnitude of change across them. Similarly, across the three THI subscales (Functional, Emotional, and Catastrophic), substantial improvements and large effect sizes were observed. Together, these outcomes from two validated and strongly correlated questionnaires highlight Lenire’s efficacy in addressing the complex and multifaceted burden of tinnitus.

The results of this study are further supported by the demonstrated safety and acceptability of the treatment. Previous clinical trials (Conlon et al. 2020, 2022; Boedts et al. 2024) and RWE studies (Buechner et al. 2022; McMahan and Lim 2025) have consistently shown that Lenire is inherently safe, with no procedure or device-related serious adverse events reported. Additionally, the recent FDA clinical trial (Boedts et al. 2024) found that any non-serious adverse events were resolved with minimal intervention, underscoring the reversible and non-consequential nature of these events and the very low risk profile associated with bimodal stimulation using Lenire. In the current RWE analysis of tinnitus patients treated at T&B, no field safety reports necessitating submission to the FDA were required, nor were any corrective actions, product removals, market withdrawal, or recalls necessary.

Given the real-world design of the current study, some follow-up data were missing, as not all patients completed the TFI and THI at FU1 or FU2. To address any potential bias arising from missing data, we employed a Markov chain Monte Carlo multiple imputation method. The imputed data analysis revealed no significant differences compared to the primary outcomes obtained from the complete case cohort. These findings suggest that non-attendance at the follow-up appointments does not necessarily reflect a negative treatment outcome but may instead indicate early clinical benefit and a reduced desire for further clinical support.

RWE studies play a vital role in evaluating how medical devices perform in everyday clinical practice. However, they inherently face certain limitations. Unlike controlled clinical trials, retrospective chart reviews rely solely on data documented during routine care, which may be incomplete or inconsistent. This can restrict the availability of detailed patient information, such as prior treatments before attending the T&B clinic, as well as important demographic and tinnitus characteristics that could aid in identifying distinct patient subgroups. Moreover, the retrospective nature of RWE studies means that no control group is available, limiting the ability to fully assess treatment effects across subpopulations or to determine optimal treatment strategies. Despite these constraints, RWE remains essential for understanding device utilization and effectiveness in real-world settings.

It is noteworthy that all patients received education and counselling at initial assessment prior to starting the Lenire treatment, which may have contributed to some therapeutic benefit. However, previous studies of counselling alone report minimal THI improvements, ranging from 1 to 3 points improvement over 12 weeks to 6 months from intervention (Cima et al. 2012; Henry et al. 2016). These findings suggest that substantial improvements observed in this study are unlikely to be attributable to counselling and education alone.

While controlled clinical trials remain the gold standard for establishing treatment efficacy, their findings do not always directly translate to real-world clinical practice. The real-world results presented in this study demonstrate that Lenire can be successfully implemented within routine audiology care, yielding significant clinical improvements as measured by both the TFI and THI. These findings complement other real-world studies that have similarly reported clinically meaningful benefits on the THI, collectively reinforcing Lenire’s effectiveness in diverse clinical settings. Notably, the findings underscore the value of the TFI as a robust and clinically relevant primary outcome measure for assessing treatment response with Lenire alongside the widely used THI. Continued accumulation of real-world data will be important to refine patient selection criteria and optimize treatment protocols, ultimately ensuring that patients can achieve optimal outcomes with the Lenire treatment.

## Acknowledgements

The authors of this study would like to extend their gratitude to the team at Tobias & Battite Hearing Wellness in Boston and Beverly, Massachusetts. Furthermore, the authors would like to thank Alex Kennedy, Sook Ling Leong, and Emma Meade at Neuromod Devices for their assistance with ethics submission, data validation, and analytics during this study. The authors are grateful to the IRB for their review and determination that the study meets the requirements of 45 CFR 46.104(d)(4).

## Data Availability

All data relating to the study are presented in the main body of the text and in the supplementary material. If access to the raw data is required, it can be obtained by contacting. The request will be handled within 10 days of receipt, from 3 months to 5 years post-article publication.

## Financial

The data was collected during the daily operations at Tobias & Battite Hearing Wellness, with funds for ethics fees being provided by Neuromod Devices.

