## Supplemental table and figures for "First Real-World Evidence Utilizing the Multidimensional Tinnitus Functional Index to Assess Treatment Impact with Bimodal Neuromodulation"

***Supplementary Material:***

| **Measure** | **Mean ± SD** |
| --- | --- |
| FU1 TFI | 36.52 ± 16.5 |
| FU1 TFI Imputed | 37.67 ± 16.6 |
| FU2 TFI | 33.43 ± 16.72 |
| FU2 TFI Imputed | 34.55 ± 16.8 |
| FU1 THI | 37.33 ± 18.11 |
| FU1 THI Imputed | 38.91 ± 18.34 |
| FU2 THI | 32.16 ± 14.14 |
| FU2 THI Imputed | 33.08 ± 13.98 |

*Supplementary Table 1: Multiple imputation for TFI and THI scores at FU1 and FU2.*

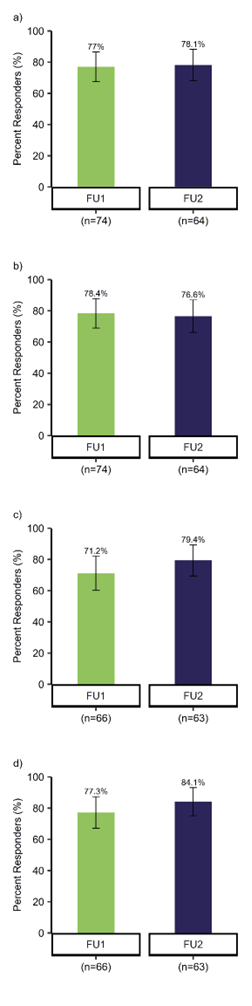

*Supplementary Figure 1: a) Percent responders using change in TFI greater than the MCID of -9 points (Engelke et al. 2025), 77.0% [95% CI: 67.4%, 86.6%] at FU1 (n=74) and 78.1% [95% CI: 68.0%, 88.3%] at FU2 (n=64). Two-sided z-test for proportions, p-value of 0.878. b) Percent responders using change in TFI greater than the MCID of 15% (Langguth and De Ridder 2023), 78.4% [95% CI: 69.0%, 87.6%] at FU1 (n=74) and 76.6% [95% CI: 66.2%, 86.9%] at FU2 (n=64). Two-sided z-test for proportions, p-value of 0.799. c) Percent responders using change in THI greater than the new MCID of -11 points (Engelke et al. 2025), 71.2% [95% CI: 60.3%, 82.1%] at FU1 (n=66) and 79.4% [95% CI: 69.4%, 89.4%] at FU2 (n=63). Two-sided z-test for proportions, p-value of 0.284. d) Percent responders using change in THI greater than the MCID of 15% (Langguth and De Ridder 2023), 77.3% [95% CI: 67.2%, 87.4%] at FU1 (n=66) and 84.1% [95% CI: 75.1%, 93.2%] at FU2 (n=63). Two-sided z-test for proportions, p-value of 0.325.*

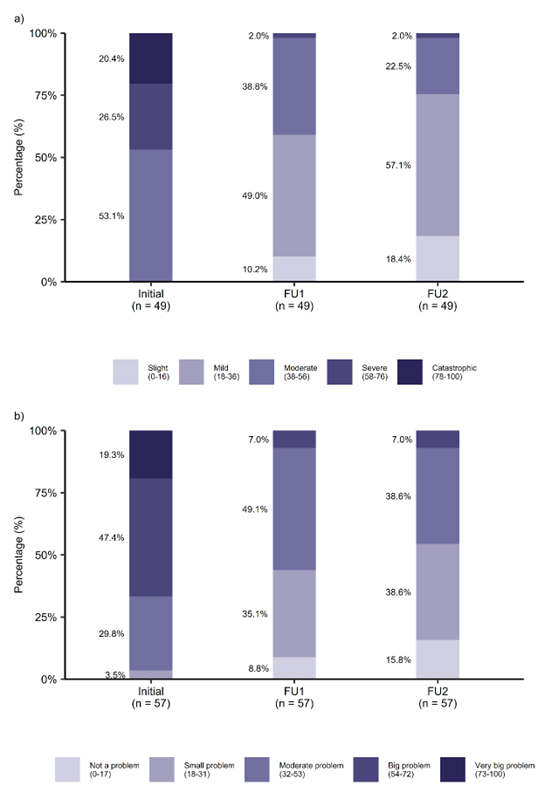

*Supplementary Figure 2: Percentage of patients with available THI (n=49) and TFI (n=57) results at all three time points (initial assessment, FU1 and FU2), categorized by severity categories at initial assessment. a) THI severity is categorized as slight (0-16 points), mild (18-36), moderate (38-56), severe (58-76), and catastrophic (78-100). b) TFI problem levels are categorized as not a problem (0-17), small problem (18-31), moderate problem (32-53), big problem (54-72), and very big problem (73-100).*
